# Bayesian spatial analysis of *Plasmodium* parasites prevalence and its determinants in selected regions of Mainland Tanzania

**DOI:** 10.64898/2026.04.13.26350696

**Authors:** Daniel P. Challe, Daniel A. Petro, Filbert Francis, Misago D. Seth, Rashid A. Madebe, Angelina J. Kisambale, Dativa Pereus, Salehe S. Mandai, Catherine Bakari, Hussein J. Semboja, Solomon Mwakasungula, Gervas A. Chacha, Rule Budodo, Daniel Mbwambo, Sijenunu Aaron, Abdallah Lusasi, Samwel Lazaro, Celina I. Mandara, Vedastus W. Makene, Deus S. Ishengoma

## Abstract

**Background:** Malaria remains a major public health challenge globally and in Tanzania, driven by persistent *Plasmodium* parasite transmission, environmental variability, and socio-economic inequalities. Despite targeted control strategies, transmission remains heterogeneous and under-captured by routine surveillance. This study utilised community cross-sectional surveys (CSS) data and spatial modelling to determine household-level risk estimates and identify micro-hotspots to guide more efficient, evidence-based malaria interventions in Mainland Tanzania.

**Methods:** The CSS data used in this study were collected in 13 villages across five regions with moderate to high malaria transmission in Mainland Tanzania between July and August 2023. Individuals aged ≥6 months, residing in the study villages for ≥3 months, were enrolled after providing informed consent and tested for malaria using rapid diagnostic tests (RDTs). Socio-demographic, clinical, anthropometric, parasitological and geo-coordinates data were collected using structured electronic tools. Household-level *Plasmodium* parasite prevalence was modelled using Bayesian geostatistical methods implemented through Integrated Nested Laplace Approximation within a Stochastic Partial Differential Equation framework, incorporating relevant environmental covariates. Model performance was evaluated using the Watanabe-Akaike Information Criterion (WAIC).

**Results:** Bayesian models with village-specific covariates consistently outperformed null models, as indicated by lower WAIC values. In Kyerwa district (Kagera region), grass cover increased the risk of *Plasmodium* parasite prevalence (Posterior mean (PM)=0.076; 95% credible interval [CrI]: 0.040–0.112), while altitude had a protective effect (PM=-0.002; 95%CrI: -0.003 to -0.001), with strong sub-village clustering of malaria infection (variance=0.485; 95% CrI [0.333 - 0.730]). In Buhigwe district (Kigoma region), shrub cover increased the risk of *Plasmodium* parasite prevalence (PM=0.119; 95% CrI: 0.029–0.210) while in Ludewa (Njombe), both shrub (PM=0.512; 95% CrI: 0.066–0.989) and grass (PM=0.490; 95% CrI: 0.117–0.879) increased the risk of infection, with pronounced sub-village clustering (variance=0.84; 95% CrI: [0.38 – 2.40]). In Nyasa district (Ruvuma), shrub cover had a modest positive effect (PM=0.070; 95% CrI: 0.005–0.135), in Muheza district (Tanga region), its effect was influential (PM=0.160; 95% CrI: 0.056–0.266). Risk maps revealed fine-scale heterogeneity in the household-level risk of *Plasmodium* parasite prevalence.

**Conclusion:** There was pronounced micro-scale heterogeneity in *Plasmodium* transmission across the study districts, driven by localised environmental factors and strong spatial dependence. Altitude had a protective effect, while vegetation cover increased the risk of infection. Geostatistical models effectively identified household-level hotspots, highlighting the limitations of aggregated surveillance, emphasising the need for locally precision-guided malaria control strategies to improve intervention efficiency and enhance the ongoing elimination strategies.

## Background

Malaria continues to be one of the most pressing public health problems worldwide, despite substantial progress made in reducing its burden over the past two decades (African Union, 2023). The disease is caused by protozoan parasites of the genus *Plasmodium* and transmitted through the bites of infected *Anopheles* mosquitoes (Djihinto et al. 2022). According to the World Health Organisation (WHO), an estimated 282 million malaria cases and 610,000 deaths were reported globally in 2024, with the WHO African Region (WHO Afro) accounting for approximately 94% of cases and 95% of malaria deaths (WHO, 2025). The burden of *Plasmodium* parasite remains disproportionately high in sub-Saharan Africa, where limited access to quality health services, weak surveillance systems, and persistent transmission are major challenges to the ongoing elimination efforts (Li et al. 2024; Bashir et al. 2025). In addition, the recently reported biological threats, climate variability, population movements, and ecological changes are the main reasons for limited progress in global malaria elimination and eradication strategies (Chapoterera et al. 2025).

In Tanzania, the *Plasmodium* parasite remains a major public health challenge and is among the leading causes of morbidity and mortality, particularly among under-fives and pregnant women (Challe et al. 2025; Adam et al. 2025). The country is characterised by heterogeneous *Plasmodium* parasite transmission that varies geographically, with prevalence ranging from very high in some regions to nearly zero in others (Thawer et al. 2020; Thawer et al. 2022; Thawer et al. 2023). This heterogeneity is influenced by ecological and environmental factors such as rainfall, altitude, vegetation cover, and temperature, as well as socio-economic determinants, including housing type, access to healthcare, and use of malaria preventive measures (Gbaguidi et al. 2025; Mukabana et al. 2025). In response, the National Malaria Control Programme (NMCP) has stratified the country into four malaria transmission categories: very low, low, moderate, and high transmission (NMCP, 2020). This stratification has guided the implementation of targeted interventions, including long-lasting insecticidal nets (LLINs), indoor residual spraying (IRS) (Demissie et al. 2025), intermittent preventive treatment in pregnancy (IPTp) (Makenga et al. 2025), and prompt case management using artemisinin-based combination therapies (ACTs) (Guissou et al. 2025).

Despite these control efforts, *Plasmodium* parasite transmission persists in several parts of the country, reflecting gaps in intervention coverage and the complex interplay of biological, environmental, and social determinants (Yitageasu et al. 2025). In particular, regions located in the north-west and western parts of Tanzania (such as Kagera and Kigoma) and southern regions, including Ruvuma remain in high-transmission settings, while Tanga exhibits moderate transmission. Others, such as Njombe, have consistently reported low to very low transmission intensities (Thawer et al. 2020; Thawer et al. 2022; Thawer et al. 2023: SMPS: 2025). Understanding the local drivers of transmission is therefore crucial for tailoring interventions that are both efficient and sustainable. Traditional surveillance systems based mainly on routine health facility data are limited in their ability to capture the true burden of *Plasmodium* malaria parasites, as they often miss asymptomatic infections and do not adequately represent rural populations where transmission is highest (Mazigo et al. 2025). Community cross-sectional surveys (CSS) have increasingly been used to address this limitation, providing reliable prevalence estimates and allowing integration of socio-demographic, clinical, and ecological data for detailed epidemiological analysis (Challe et al. 2025; Chacha et al. 2025; Mandai et al. 2024; Challe et al. 2025).

In recent years, advances in geospatial modelling and Bayesian statistical approaches have provided powerful tools for analysing *Plasmodium* malaria parasites transmission dynamics. Spatial statistical methods, particularly those based on Bayesian inference, enable the estimation of disease risk at fine geographic scales, accounting for uncertainty and spatial correlation (Mahama 2022). The Integrated Nested Laplace Approximation (INLA) framework and the stochastic partial differential equation (SPDE) approach are among the most widely applied methods, allowing for flexible and computationally efficient modelling of spatial processes (Zheng et al. 2025). These methods are particularly well-suited to malaria research; this study used these Bayesian spatial approaches to determine Plasmodium parasite prevalence and risk factors in selected regions of Mainland Tanzania.

## Methods

### Study design and site

The study used data from a CSS conducted between July and August 2023 in 13 villages across five regions of Mainland Tanzania: Kagera, Kigoma, Njombe, Ruvuma, and Tanga (Fig.1). The surveys were carried out during or shortly after the peak of malaria transmission season and were part of the project on Molecular Surveillance of Malaria in Mainland Tanzania (MSMT)., The MSMT project aimed at building the comprehensive capacity for integrated malaria molecular surveillance (MMS, covering both parasites and vectors), and it was implemented in 13 regions in 2021–2022, with further expansion to cover all 26 regions of Mainland Tanzania in 2023, as previously reported (Popkin-Hall et al. 2024; Popkin-Hall et al. 2024; Derua et al. 2025).

**Figure 1:**
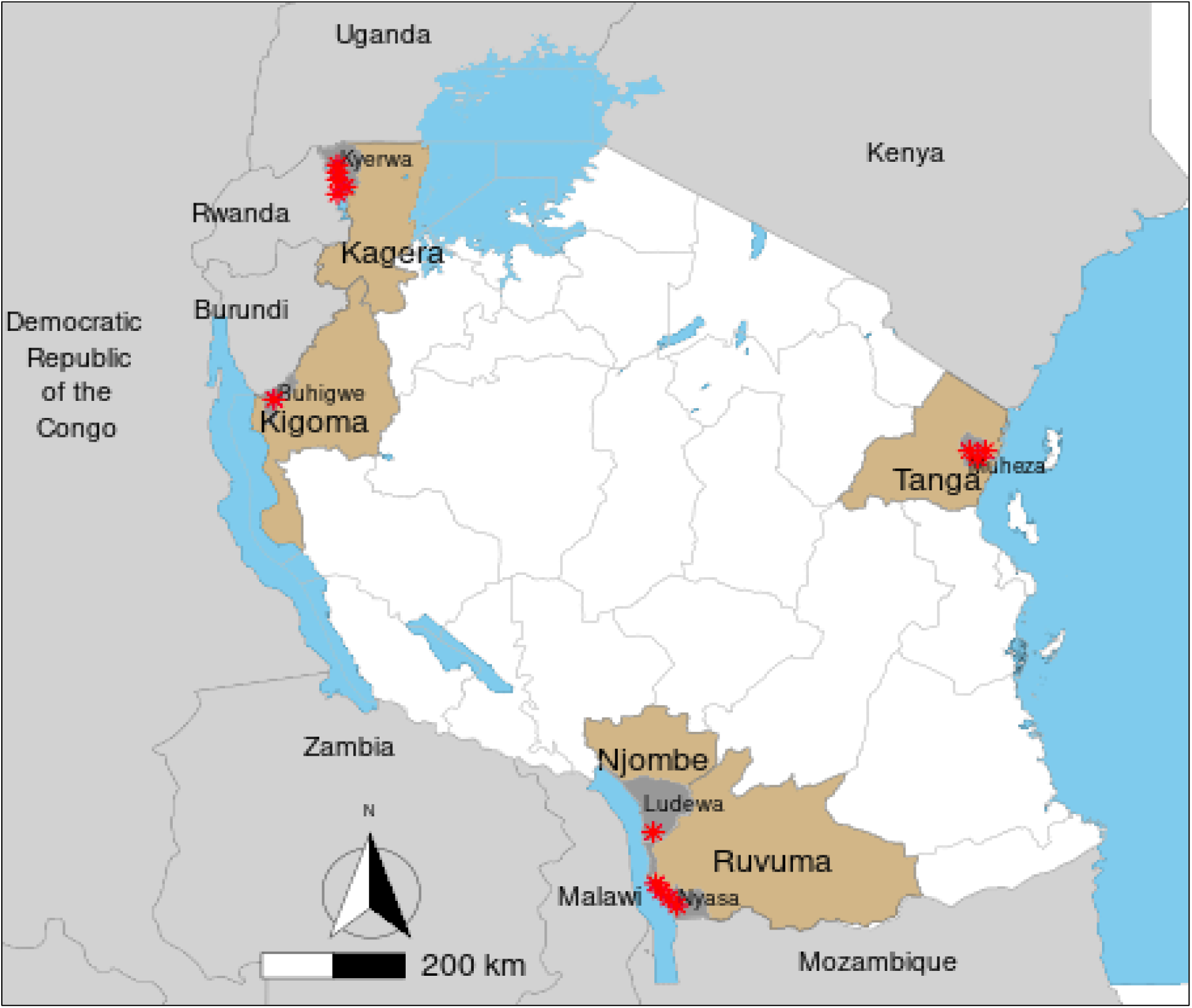
Map of Tanzania showing the study regions (grey), study districts (light blue) and study villages (red stars)

The selected regions represented varying malaria transmission intensities, with Kagera, Kigoma and Ruvuma classified as high-transmission areas, Tanga as a moderate-transmission area, and Njombe as a very low-transmission area, according to the NMCP’s 2020 malaria stratification (Thawer et al. 2020; Thawer et al. 2022; Thawer et al. 2023). In this stratification, regions were grouped into four categories: high, moderate, low, and very low transmission. The study initially focused on communities which were previously studied by the authors (Challe et al. 2025; Chacha et al. 2025; Challe et al. 2025; Mandai et al. 2024; Ishengoma et al. 2018; Francis et al. 2017; Liheluka et al. 2023; Ishengoma et al. 2024; Bakari et al. 2020; Ishengoma et al. 2011; Seth et al. 2025; Budodo et al. 2025), thereby building on extensive historical data to strengthen analyses of malaria trends using both new and existing information. The two regions of Kagera and Njombe were included in the CSS in 2023 due to reports of confirmed artemisinin partial resistance in Kagera (Ishengoma et al. 2024; Juliano et al. 2024) and sporadic occurrence of histidine-rich protein 2/3 (hrp2/3) gene deletions in both Kagera and Njombe (Rogier et al. 2024).

In Kagera, the study villages were Kitwechenkura, Nyakabwera, Rubuga, Kitoma, and Ruko in Kyerwa district, bordering Rwanda, where ART-R has been recently confirmed (Ishengoma et al. 2024; Juliano et al. 2024; Mandai et al. 2024). In Kigoma, Nyankoronko village (Buhigwe district), located near the Burundi border, was included in the CSS because it has been studied by the same authors from 2017 (Challe et al. 2025; Liheluka et al. 2023). In Ruvuma, three villages from Nyasa district (Lundo, Lipingo, and Ngindo) situated along the shores of Lake Nyasa bordering Malawi were studied, because they were also included in other studies before and after the launch of the MSMT project as previously described (Challe et al. 2025; Chacha et al. 2025). From the Njombe region, Kipangala village (Ludewa district), also on the shores of Lake Nyasa near the Malawi border, was included because it has the highest prevalence of single *hrp2* gene deletions compared to other districts covered in the nationwide *hrp2/3* gene deletions conducted in 2021 (Chacha et al. 2025; Rogier et al. 2024). In the Tanga region, the villages of Magoda, Mamboleo, and Mpapayu (Muheza district) were selected, owing to their long history of involvement in malaria research since 1992, as reported elsewhere (Challe et al. 2025; Chacha et al. 2025; Challe et al. 2025; Ishengoma et al. 2018; Francis et al. 2017; Ishengoma et al. 2011). Several of these areas, including Muheza district in Tanga, have a long history of malaria research, providing a strong foundation for longitudinal analyses of transmission trends (Ishengoma et al. 2018; Ishengoma et al. 2013; Francis et al. 2017). Importantly, the study villages are located in ecologically diverse environments: from highland border districts such as Kyerwa in Kagera (19,21) to lowland lake shore villages in Ruvuma and Njombe (Liheluka et al. 2023; Ishengoma et al. 2024). These variations provide a natural laboratory for examining how altitude, vegetation density, and other environmental factors influence the risk of infection by *Plasmodium* malaria parasites

### Study population and recruitment of participants

This study targeted and recruited individuals aged ≥6 months residing in the selected villages. The study targeted registered individuals from each village, based on census surveys that were conducted before each round of CSS as described previously (Mandai et al. 2024; Chacha et al. 2025; Challe et al. 2025; Challe et al. 2025). Eligibility criteria included residence in the study villages, age ≥6 months, and provision of informed consent or assent as previously described (Challe et al. 2025; Chacha et al. 2025; Challe et al. 2025; Mandai et al. 2024). Individuals from the villages that were not under the MSMT project or those who did not consent were excluded. Before the CSS, the project team collaborated closely with village leaders to develop the survey schedules in each village. All community members were informed about the surveys and were invited to participate voluntarily, and this was done by community sensitisation teams, which were under the leadership of village authorities. The sensitisation and invitation of participants to participate in the CSS were done using loudspeakers and other local communication channels. The announcements included details on survey objectives, dates for the respective villages, and participation procedures. The schedules for each village ensured adequate coverage of the village based on the number of sub-villages (hamlets) and the total village population. To enhance accessibility, CSS posts were established in centrally located hamlets to maximise reach. Where necessary, the CSS posts were rotated among hamlets to ensure equitable access across the village. Participants were mobilised and invited according to their hamlet of residence, and recruitment was conducted at convenient locations. Participation in the CSS was entirely voluntary, and community members who were unable to attend on their designated day were offered alternative opportunities to participate, either on another day within the same village or in a nearby village (particularly in Kagera, Ruvuma and Tanga, where more than one village was involved in the CSS). Data collection in each village was completed within 2–5 days, depending on village population size, and the team ensured that the required sample size was achieved, as previously reported (Challe et al. 2025; Chacha et al. 2025; Challe et al. 2025; Mandai et al. 2024).

### Data collection procedures

The CSS were preceded by census surveys, which were carried out in each village using structured questionnaires. These censuses aimed to enumerate and register all households, and collect individual-level data for every family member. Each household was properly identified and assigned a unique identification (ID) number as previously described (Mandai et al. 2024; Chacha et al. 2025; Challe et al. 2025; Challe et al. 2025). Household information, individual data and geo-coordinates were updated each annual census. New households and members were incorporated into the project database accordingly.

During the CSS, participants were identified using their unique IDs assigned during census surveys. These IDs were generated from the project database prior to each survey. Any individual missing from the pre-prepared CSS list was added to the census register and invited to participate. Data collection was conducted by trained and experienced project staff, supported by community health workers (CHWs). The CHWs were recruited from the same villages, had participated in previous surveys, and received training or refresher training when necessary. The data were collected using structured questionnaires incorporated into the project’s case report forms (CRFs). The CRFs were completed by experienced MSMT project staff who had participated in previous CSS or were trained/retrained before the survey.

The data collection process involved verifying and updating demographic information from the project database and collecting anthropometric, parasitological, and clinical data as previously described (Mandai et al. 2024; Chacha et al. 2025; Challe et al. 2025; Challe et al. 2025). Each eligible participant was assigned a survey-specific ID and issued a registration card containing the required personal information. After registration, participants were interviewed to obtain socio-demographic data and information on malaria prevention practices.

Participants then proceeded to the anthropometric section, where body weight, height, and axillary temperature were measured by the research team with assistance from CHWs. Thereafter, they moved to the laboratory section, where finger-prick blood samples were collected for the detection of malaria parasites using rapid diagnosis tests (RDTs). The RDTs used in the CSS included Abbott Bioline Malaria Ag Pf/Pan (Abbott Diagnostics Korea Inc., Gyeonggi-do, South Korea) and Malaria Pf/Pan Ag Rapid Test Cassette (Zhejiang Orient Gene Biotech Co., Ltd., Zhejiang, China). From the same finger prick, dried blood spots (DBS) on filter papers (Whatman No. 3, GE Healthcare Life Sciences, PA, USA) and both thin and thick blood smears were collected for further laboratory analysis. Some laboratory findings have been reported elsewhere (Mandai et al. 2024; Chacha et al. 2025; Challe et al. 2025; Challe et al. 2025).

Finally, participants proceeded to the clinical section, where study clinicians conducted clinical assessment, including documentation of any illness history and treatments taken within the seven days preceding the survey. A physical examination was performed, and clinical diagnoses were made accordingly. Participants who tested positive for malaria were treated according to the national treatment guidelines (NMCP. 2020), while those diagnosed with other illnesses received appropriate management based on their clinical condition (MoH. 2017).

The CSS workflow was optimised by the MSMT project to ensure smooth, one-directional movement of participants from registration through laboratory testing to the clinical section. Participants arrived at the clinical section with their RDT results, thereby preventing unnecessary back-and-forth movement. This design eliminated the need for repeat clinical visits that would otherwise occur if the clinical section were positioned before the laboratory section, as described earlier (Mandai et al. 2024; Chacha et al. 2025; Challe et al. 2025; Challe et al. 2025).

### Data extraction and management

All census and CSS data were collected using the MSMT project’s CRFs, which were optimised to run on the Open Data Kit (ODK) software installed and run on tablets. The data were collected offline; however, once the study team gained internet access, they transferred the data for storage on a central server located at NIMR in Dar es Salaam. Thereafter, the data were downloaded into Excel spreadsheets for cleaning daily, and after the completion of the surveys. Additional cleaning was done using Excel and STATA version 17 (STATA Corp Inc., TX, USA). Vegetation cover for 2023 was extracted from Google Earth Engine (https://earthengine.google.com/) using village shapefiles. All datasets were then exported to Excel and verified for completeness before risk modelling and epidemiological analysis. Covariates, including vegetation cover (grass and/or shrub) and altitude, were included as fixed effects in the model.

### Statistical analysis

In the first step, data with their geocoordinates were aggregated at the household level to investigate clustering of the relative risk of *Plasmodium* parasites among individuals who participated in the selected study villages. The outcome was the number of individuals testing positive for *Plasmodium* parasites by RDTs out of the total number examined per household. Given that each household was represented as a point, household-level risk of *Plasmodium* parasite prevalence was modelled using a Bayesian geostatistical framework implemented via Integrated Nested Laplace Approximation (INLA) (Mougeni et al. 2024). A binomial likelihood with a logit link was used to model household-level prevalence, with spatial dependence captured by a latent Matérn Gaussian random field. The spatial field was represented using the Stochastic Partial Differential Equation (SPDE) approach, which approximates the continuous Matérn field as a Gaussian Markov random field defined over a triangulated mesh. Household coordinates were linked to the mesh through the SPDE projection matrix (Roques et al. 2021), enabling spatial interpolation and prediction (Mougeni et al. 2024).

Two hierarchical models were fitted. The first model (null model [without covariates]) included an intercept, a spatial random field, and a random intercept only for the village and sub-village to capture hierarchical clustering. The second model (with covariates) extended this structure by adding environmental covariates (altitude, grass cover and shrub cover) as fixed effects (Damien et al. 2022). Village and sub-village effects were modelled as independent and identically distributed Gaussian random intercepts. All models were fitted using INLA, with full computation of posterior marginals for fixed effects, random effects and hyperparameters (Bivand et al. 2015).

Model comparison was based on the Watanabe-Akaike Information Criterion (WAIC), while the Deviance Information Criterion (DIC) was computed to assess model fit and effective complexity (Du et al. 2024). For validation, the dataset was randomly partitioned into training (80%) and testing subsets, and predictive accuracy was assessed on the held-out data. Posterior summaries of the spatial field, including the marginal variance and nominal range, were extracted and presented as texts. Random effect precisions were converted to variance to quantify village and sub-village-level heterogeneity. Covariates for the final model in each village were selected based on two criteria: a significant association with the outcome variable or an improvement in the model’s WAIC. Consequently, the specific set of covariates varied across study villages. To determine the most robust approach, a comparison of the null model (Model 1, without covariates) and the model with the selected covariates (Model 2) was conducted using WAIC. Lower DIC and WAIC scores indicate superior predictive quality and model fit, so Model 2, which consistently yielded lower values, was selected as the most suitable for risk modelling. Results are reported as posterior means (PM), while PM >0 indicates risk and PM<0 shows protection with their corresponding 95% credible intervals (CrI). To visualise spatial patterns, maps of predicted *Plasmodium* parasite risk were generated across each study village. All statistical analyses were performed using R software version 4.3.2 via the R-INLA package.

## Results

### Baseline characteristics of participants and details of the data

A total of 8,326 individuals from 1,243 households in 13 villages were included in the study. Kyerwa contributed the largest proportion of participants, with 4,001 (48.1%) individuals, 440 (35.4%) households and 5 (38.5%) villages. Nyasa was the second largest contributor, with 1,891 (22.7%) individuals and 270 (21.7%) households across three villages. Muheza had 1,231 (14.8%) individuals from 88 (7.1%) households in three villages. Each of the two districts of Buhigwe and Ludewa had only one village and contributed similar proportions of individuals, with 609 (7.3%, from 238 households) and 594 (7.1%, from 207 households) individuals, respectively. In the five districts, altitude varied markedly, ranging overall from 139.7 to 1,694.4 metres above sea level. Kyerwa and Buhigwe were predominantly highland districts, while Nyasa and Muheza were mainly lowland settings (Table 1).

**Table 1:** Baseline characteristics of participants and details of the data.

| Variable | Districts |  |  |  |  | Total<br>n (%) |
| --- | --- | --- | --- | --- | --- | --- |
|  | Kyerwa<br>n (%) | Buhigwe<br>n (%) | Ludewa<br>n (%) | Nyasa<br>n (%) | Muheza<br>n (%) |  |
| Individuals | 4,001 (48.1) | 609 (7.3) | 594 (7.1) | 1,891<br>(22.7) | 1,231<br>(14.8) | 8,326 (100.0) |
| Households | 440 (35.4) | 238 (19.1) | 207 (16.7) | 270 (21.7) | 88 (7.1) | 1,243 (100.0) |
| Number of<br>villages | 5 (38.5) | 1 (7.7) | 1 (7.7) | 3 (23.1) | 3 (23.1) | 13 (100.0) |
| Altitudes range<br>in metres (min -<br>max) | 1,065.5 –<br>1,694.4 | 1,200.9 –<br>1359.8 | 713.6 –<br>1,334.2 | 252.0 –<br>676.7 | 139.7 –<br>461.9 | 139.7 –<br>1,694.4 |

### Spatial variation of the risk of *Plasmodium* parasites prevalence

The study covered a wide geographic distribution of sampled households across study villages, and demonstrated spatial coverage of the study population in Kyerwa, Buhigwe, Ludewa, Nyasa and Muheza districts (Figure 2A, 2B, 2C, 2D and 2E). The findings revealed significant geographic variations in *Plasmodium* parasite prevalence across the study villages (Figure 3A, 3B, 3C, 3D and 3E). In Kyerwa district, vegetation cover (grass) was positively associated with the risk of malaria infection (PM = 0.076; 95% CrI: 0.040 – 0.112), while altitude showed a protective effect (PM = –0.002; 95% CrI: -0.003 to -0.001). Furthermore, village-level variation was small (0.045; 95% CrI: 0.011–0.452) compared to sub-village, which was considerably higher (0.485; 95% CrI: 0.333 0.730) (Table 2). This disparity indicates substantial clustering of transmission at the sub-village level, driven primarily by localised conditions. In Buhigwe, only vegetation cover (shrub) was positively associated with the risk of Plasmodium parasite infection (PM = 0.119; 95% CrI: 0.029 – 0.210) with no village and sub-village level variation (Table 2). In Ludewa district (Table 2), both grass (PM = 0.490; 95% CrI: 0.117 – 0.879) and shrub (PM = 0.512; 95% CrI: 0.066–0.989) were positively associated with the risk of *Plasmodium* parasite infection. On the other hand, altitude was negatively associated with the outcome (PM = -0.015; 95% CrI: -0.022 to -0.009), indicating lower risk in households with higher elevations. Village-level variation was extremely small (0.000046; 95% CrI; 0.000012 – 0.000652), indicating negligible heterogeneity between villages. In contrast, sub-village variation in the risk of *Plasmodium* parasite infection was high (0.84; 95% CrI: 0.38 – 2.40), suggesting substantial clustering at the sub-village scale. This indicates that fine-scale local factors, rather than broader village-level differences, explain most of the observed heterogeneity in the risk of Plasmodium parasite transmission. For Nyasa district (Table 2), only vegetation cover (shrub) (PM = 0.070; 95% CrI: 0.005 – 0.135) was positively associated with *Plasmodium* parasite infection. Village-level variation in the risk of Plasmodium parasite infection was extremely small (0.000045; 95% CrI: 0.000012 – 0.000665), compared to sub-village level (0.0275; 95% CrI: 0.0069 – 0.160). This suggests modest clustering of the Plasmodium parasite infection at the sub-village level. The result from Muheza district (Table 2) shows that shrubs were positively associated with *Plasmodium* parasite infection (PM = 0.160; 95% CrI: 0.056 – 0.266), indicating higher malaria risk in areas with more vegetation cover. Altitude showed a weak negative association (PM = –0.003; 95% CrI: -0.006 – 0.001) while village- (0.000036; 95% CrI: 0.000009 – 0.000441) and sub-village level (0.000036; 95% CrI: 0.000009 – 0.000414) variations were extremely small. This indicates that administrative-level clustering contributed almost no additional variation once environmental covariates were included.

**Figure 2:**
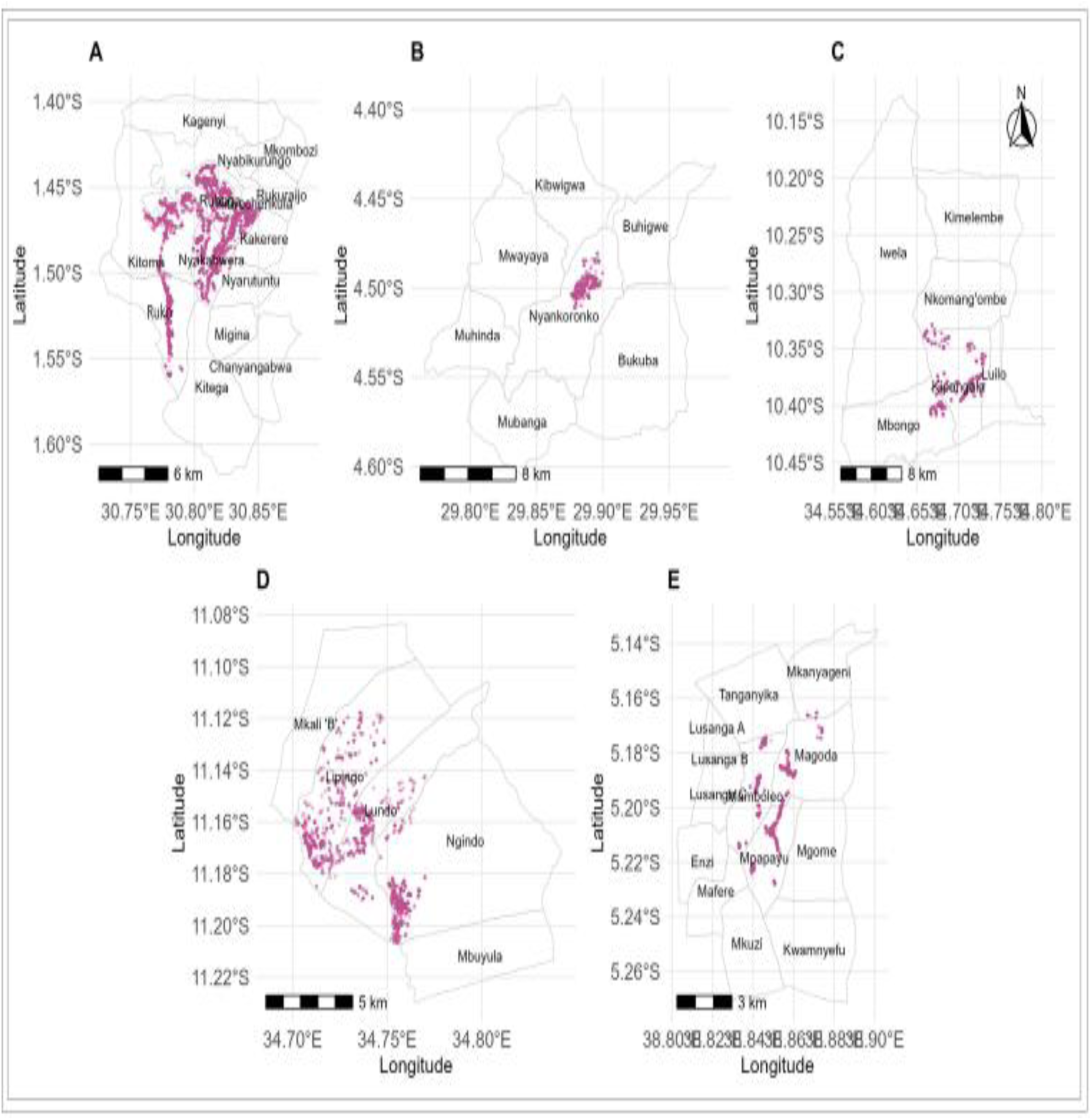
Households distribution (red dots) in the study villages; A: Villages in Kyerwa district, B: Village in Buhigwe, C: Village in Ludewa, D: Villages in Nyasa and E: Villages in Muheza districts.

**Figure 3:**
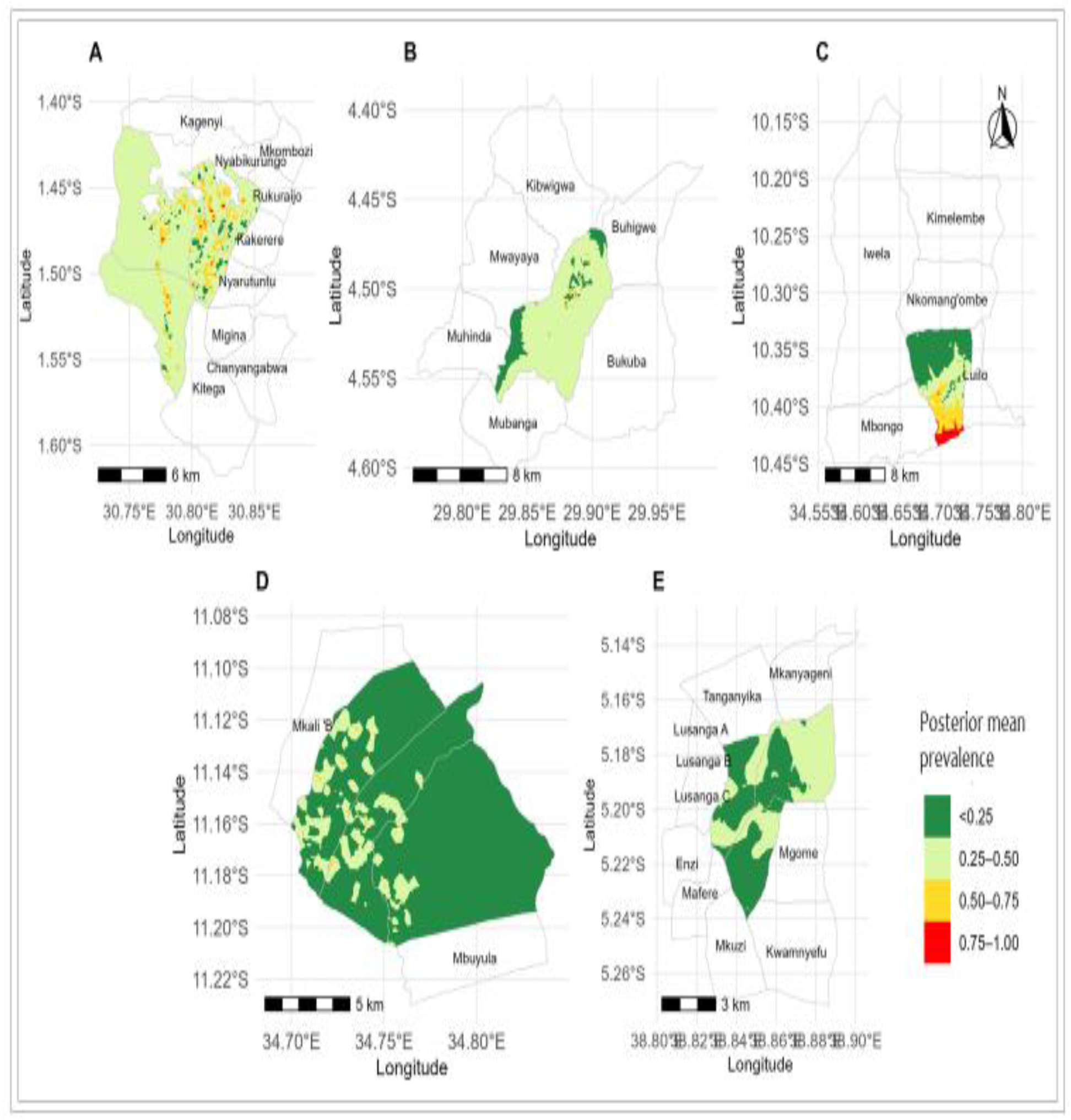
Smooth maps showing predicted PM probability of *Plasmodium* parasite prevalence at the household level, adjusted for various covariates. A: Villages in Kyerwa district, B: Village in Buhigwe, C: Village in Ludewa, D: Villages in Nyasa and E: Villages in Muheza district.

**Table 2.**
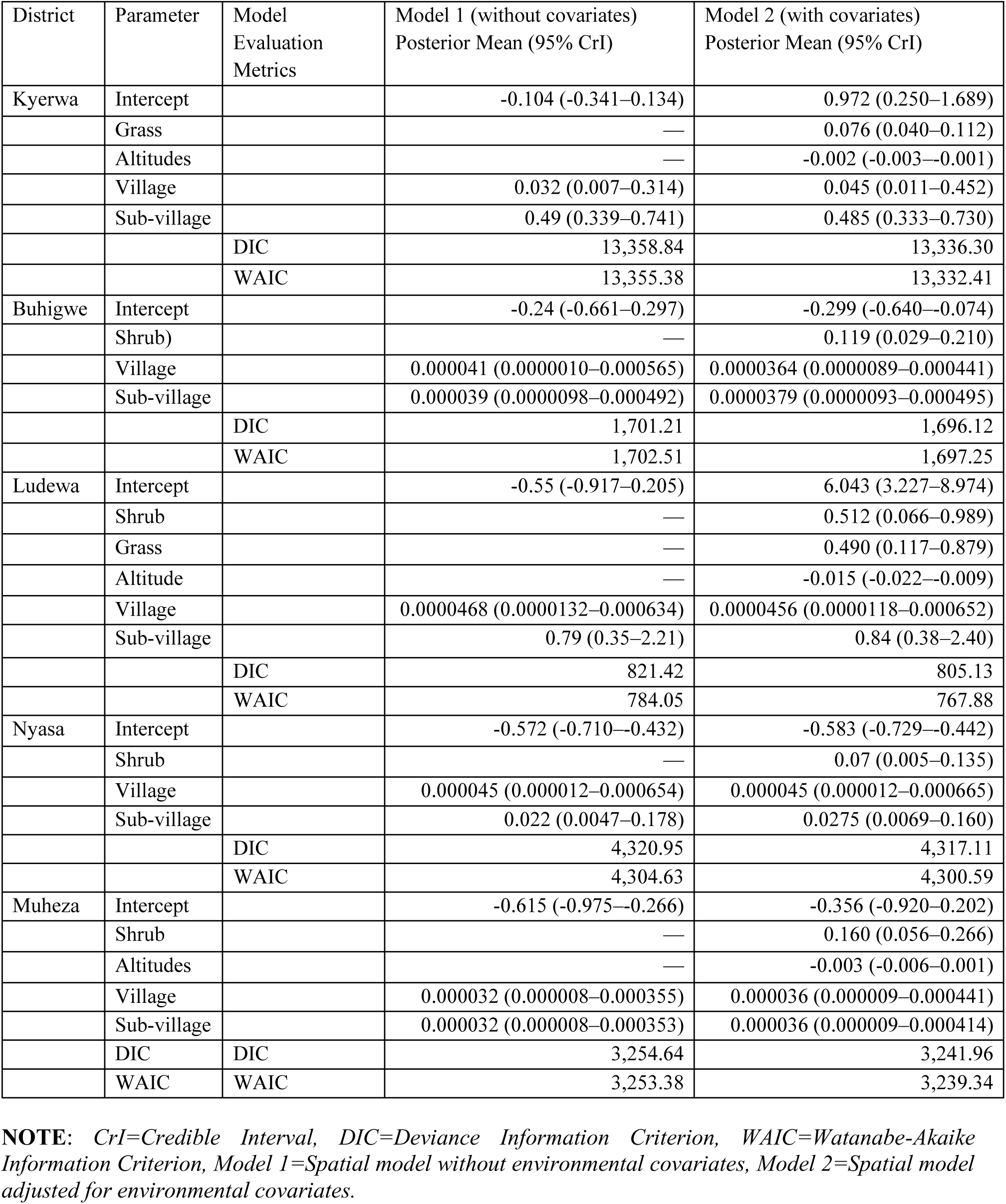
Posterior means (95% CrI) for district-specific parameters and model fit criteria.

## Discussion

This study provides detailed evidence of fine-scale spatial heterogeneity in the *Plasmodium* parasite transmission across five districts in Tanzania, illustrating how environmental factors and spatial dependence jointly shape malaria risk. Using Bayesian spatial models with a stochastic partial differential equation (SPDE) framework, the results highlight marked geographic variations in both environmental associates and spatial clustering of infections. These findings reinforce the critical importance of localised malaria epidemiology for achieving targeted and effective control interventions.

Across several districts, altitude emerged as a consistent protective factor, particularly in Kyerwa and Ludewa, where higher elevations significantly reduced household-level parasite prevalence. These results are biologically plausible and align with previous studies showing that cooler temperatures at higher altitudes limit vector survival and slow *Plasmodium* parasite development, thereby lowering transmission potential (Zong et al. 2024). Although Nyasa exhibited a similar negative trend, the association was weak and statistically non-significant, suggesting that altitude alone may not fully explain the *Plasmodium* parasite transmission ecology in this setting. Differences in microclimate, seasonal temperature variation, settlement patterns, and vector species composition may contribute to the spatial variability in altitude effect across districts (Mayilsamy et al. 2025; Nyasa et al. 2022).

In contrast, vegetation demonstrated strong but site-specific associations with malaria prevalence. Grass cover in Kyerwa and shrub cover in both Buhigwe and Muheza were positively and significantly associated with infection risk, suggesting that particular vegetation structures create microhabitats favourable for vector breeding, resting, or survival (Kahamba et al. 2024; Dzokoto et al. 2023). Grasslands and shrub-dominated landscapes often indicate proximity to water bodies, agricultural zones, or humid ecotones that support *Anopheles* larvae and adult resting sites (Dida et al. 2018). The absence of significant vegetation effects in Nyasa may indicate that other environmental or anthropogenic factors, such as agricultural practices, domestic water storage, housing structure, or vector control operations, play a more dominant role in shaping *Plasmodium* parasites transmission patterns (M.Palaniyandi 2021; Gbaguidi et al. 2025).

Spatial maps provided complementary insights by identifying clear micro-hotspots within each district, even after adjusting for environmental covariates (Iddrisu et al. 2024; Monteiro et al. 2025). The strong spatial autocorrelation captured by the SPDE random field underscores the importance of unmeasured but spatially structured determinants such as soil moisture, vector breeding micro-sites, housing density, or human mobility patterns (Zegeye et al. 2025). These *Plasmodium* parasite hotspots represent critical locations for targeted control and highlight the need for precision surveillance systems capable of detecting persistent focal transmission (Stresman et al. 2019).

Notably, model performance varied across districts. The lower DIC and WAIC values observed in Ludewa and Buhigwe indicate more parsimonious and well-fitting models. In contrast, higher values in Kyerwa suggest underlying complexity or substantial unmeasured heterogeneity of *Plasmodium* parasites in that area (Bonza et al. 2025). Differences in environmental gradients, ecological diversity, and vector behaviour likely contribute to the varying model fits. Nonetheless, the SPDE-based models effectively captured the non-linear and spatially correlated risk patterns across all sites, demonstrating the added value of spatially explicit Bayesian approaches for understanding *Plasmodium* parasites ecology (Alahmadi and Moraga 2025; Anderson et al. 2022). These findings carry important implications for *Plasmodium* parasite control. Firstly, the marked heterogeneity across districts supports the need for geographically planned intervention strategies. Areas where vegetation plays a prominent role, such as Kyerwa, Buhigwe, and Muheza, may benefit from targeted larval source management, environmental modification, or improved agricultural water governance (Otambo et al. 2022; Msugupakulya et al. 2024; Oyibo et al. 2021). Highland communities with naturally reduced risk of *Plasmodium* parasites may require more differentiated resource allocation, focusing efforts on lower-elevation valleys or fringe zones where risk clusters persist (Mwangungulu et al. 2023; Wanjala and Kweka 2016). Secondly, the study illustrates the potential of high-resolution spatial modelling for informing focal IRS, enhanced LLIN distribution, or community-based environmental sanitation campaigns. Such precision-targeted approaches may improve cost-effectiveness and accelerate progress towards malaria elimination in diverse ecological settings (Borkovska et al. 2022; Nkya 2026; Kelly et al. 2011).

This study has limitations. The cross-sectional design precludes causal inference and does not account for seasonality or temporal shifts in environmental conditions. Some environmental variables, especially vegetation indices derived from remote sensing, may be affected by temporal mismatches or classification inaccuracies. Furthermore, household-level behavioural and structural factors such as LLIN use, indoor residual spraying, occupation, house construction, and water storage were not included and may confound the observed associations. Despite these limitations, the study provided a robust spatial assessment of micro-level malaria heterogeneity across ecologically distinct settings.

## Conclusion

The study demonstrated a significant geographic variability in the environmental and spatial determinants of malaria transmission across Tanzania. By using advanced Bayesian spatial methods, it highlights the complex, location-specific drivers of the *Plasmodium* parasite prevalence and emphasises the value of precision-oriented surveillance and intervention strategies. Future research incorporating longitudinal data, entomological surveys, and additional human behavioural factors will be essential to fully elucidate the pathways underlying observed spatial patterns and to guide targeted malaria control efforts in heterogeneous environments.

## Data Availability

The data used for this study are available upon a reasonable request from the corresponding author, with institutional approval by NIMR and after signing a data transfer agreement (DTA) between NIMR and the recipient.

## List of abbreviations

ACT: Artemisinin-based combination therapy
CHWs: Community Health Workers
CRFs: Case Report Forms
CrI: Credible Intervals
CSS: Cross-sectional survey
DIC: Deviance Information Criterion
INLA: Integrated Nested Laplace Approximation
IRS: Indoor residual spraying
ITNs: Insecticide-treated bed nets
MSMT: Molecular Surveillance of Malaria in Mainland Tanzania
NIMR: National Institute for Medical Research
NMCP: National Malaria Control Program
ODK: Open Data Kit software
PM: Posterior Means
RDT: Rapid Diagnostic Test
PO-RALG: President’s Office, Regional Administration and Local Government
SPDE: Stochastic Partial Differential Equation
SSA: Sub-Saharan Africa
WAIC: Watanabe-Akaike Information Criterion
WHO: World Health Organisation

## Declarations

### Ethical approval and consent to participate

The ethical approval to conduct this study was obtained from the Medical Research Coordinating Committee of the National Institute for Medical Research (NIMR) in Tanzania, with ethical clearance number Ref. NIMR/HQ/R.8a/Vol.1/2193 dated 15^th^ November 2022. Permission to conduct the study in the villages was provided by the President’s Office, the Regional Administration and Local Government Authority, and health authorities of Tanga, Kigoma, and Ruvuma regions and districts, as well as village authorities. Informed consent/assent was sought and obtained before conducting the study from each participant or the parents/legal guardians of children. Permission to publish this paper was sought and given by the Director General of NIMR.

### Consent for publication

Not applicable.

### Author’s contributions

DPC, VWM and DSI conceptualised and designed the study and supervised data collection. All authors participated in data collection under the supervision of DSI and VWM. DPC, DAP and FF managed and analysed the data. DPC and DSI wrote the manuscript with the support of VWM and DAP, FF, MDS, RAM, AJK, DP, SSM, CB, HJS, SM, GAC, RB, DM, SA, AL, SL, CIM and made critical revisions to the initial draft and the final version. All authors read and approved the final version of the manuscript.

## Acknowledgments

The authors wish to thank all study participants for their readiness to participate and take part in this study. They highly appreciate the support of village leaders, districts, and regional medical authorities to the study team during the entire study period. Thanks to the efforts of a highly committed study team, which implemented the different aspects of the study; they include Ezekiel Malecela, Muhidin Kassim, Juma Tupa, Gineson Nkya, Neema Barua, Francis Chambo, Sawaya Msangi, Ally Idrisa, Kusa Mchaina, Christian Msokame, Rogers Msangi, Salome Simba, Ambele Lyatanga, Tumaini Kamna, Juma Akida, Salome Simba, Tilaus Gustav, Godbless Msaki, Hatibu Athuman, Gerion Rwaibebe, Emmanuel Kessy, George Gesase, Oswald Oscar, and Ildephonce Mathias. Authors also appreciate the support of the finance, administrative, and logistic support teams at NIMR: Christopher Masaka, Millen Meena, Beatrice Mwampeta, Gracia Sanga, Neema Manumbu, Halfan Mwanga, Arison Ekoni, Twalipo Mponzi, Pendael Nasary, Denis Byakuzana, Alfred Sezary, Emmanuel Mnzava, John Samwel, Daud Mjema, Seth Nguhu, Thomas Semdoe, Sadiki Yusuph, Alex Mwakibinga, Rodrick Ulomi and Andrea Kimboi. Technical and logistics support from partners at Brown University, the University of North Carolina at Chapel Hill, and the Gates Foundation team is highly appreciated.

## References

African Union. Malaria Progress Report. 2023.

Djihinto OY, Medjigbodo AA, Gangbadja ARA, Saizonou HM, Lagnika HO, Nanmede D, et al. Malaria-Transmitting Vectors Microbiota: Overview and Interactions with Anopheles Mosquito Biology. Front Microbiol. 2022;13(891573):1–12.

WHO. World malaria report 2025: Addressing the threat of antimalarial drug resistance. 2025. 1–212 p.

Li J, Docile HJ, Fisher D, Pronyuk K, Zhao L. Current Status of Malaria Control and Elimination in Africa: Epidemiology, Diagnosis, Treatment, Progress and Challenges. J Epidemiol Glob Health. 2024;14(3):561–79.

Bashir SG, Ahmed NI, Abdullahi YB, Abdi YH, Abdi MS, Musa MK. The burden of malaria in East Africa: prevalence, risk factors, and control strategies. Malar J. 2025;24(255):1–11.

Challe DP, Petro DA, Francis F, Seth MD, Madebe RA, Mandai SS, et al. Temporal and spatial trends of the prevalence of infections caused by Plasmodium parasites among rural community members in three regions with varying transmission intensities in Mainland Tanzania. Malar J. 2025;24(301):1–20.

Adam J, Luoga P, Nyamhanga T, Makunenge C, Ayubu M. Prevalence and determinants of malaria among children aged 6 – 59 months in Tanzania: a nationwide cross-sectional study. Malar J. 2025;24(248):1–9.

Thawer S, Golumbeanu M, Lazaro S, Chacky F, Munisi K, Aaron S, et al. Spatio-temporal modelling of routine health facility data for malaria risk micro-stratification in mainland Tanzania. Sci Rep. 2023;13(10600):1–11.

Thawer SG, Golumbeanu M, Munisi K, Aaron S, Chacky F, Lazaro S, et al. The use of routine health facility data for micro - stratification of malaria risk in mainland Tanzania. Malar J. 2022;21(345):1–14. Available from: 10.1186/s12936-022-04364-7

Thawer SG, Chacky F, Runge M, Reaves E, Mandike R, Lazaro S, et al. Sub-national stratification of malaria risk in mainland Tanzania: A simplified assembly of survey and routine data. Malar J. 2020;19(177):1–12. Available from: 10.1186/s12936-020-03250-4

Gbaguidi GJ, Topanou N, Filho WL, Begedou K, Ketoh GK. Environmental and socio-economic determinants of malaria transmission in West Africa: a systematic review. One Heal Outllok. 2025;7(47):1–16.

Mukabana LN, Mshani IH, Gachohi J, Minja EG, Jackson FM, Kahamba NF, et al. Heterogeneous malaria transmission patterns in southeastern Tanzania driven by socio-economic and environmental factors. Malar J. 2025;24(172):1–12. Available from: 10.1186/s12936-025-05418-2

NMCP. National Malaria Strategic Plan 2021-2025: Transitioning to Malaria Elimination Phase. 2020.

Demissie DB, Fetensa G, Desta T, Tiyare FT. Effectiveness and Efficacy of Long-Lasting Insecticidal Nets for Malaria Control in Africa: Systematic Review and Meta-Analysis of Randomized Controlled Trials. Int J Environ Res Public Health. 2025;22(1045):1–33.

Makenga G, Mmbando B, Seth MD, Baraka V, Challe DP, Francis F, et al. Implementation and effectiveness of intermittent preventive treatment in school aged children using dihydroartemisinin- piperaquine to reduce malaria burden: an implementation research of a cluster randomised trial in Tanzania. eClinicalMedicine. 2025;90(103628):1–14. Available from: 10.1016/j.eclinm.2025.103628

Banek KE. Evaluation of Adherence to Artemisinin-based Combination Therapy for the Treatment of Uncomplicated Malaria in Sierra eone. 2019.

Yitageasu G, Worede EA, Alemu EA, Tigabie M, Birhanu A. Malaria prevalence and its determinants across 19 sub-Saharan African countries: a spatial and geographically weighted regression analysis. Malar J. 2025;24(305):1–30.

Mazigo E, Jun H, Lee W jong, Louis JM, Fitriana F, Syahada JH, et al. Prevalence of asymptomatic malaria in high- and low-transmission areas of Tanzania: The role of asymptomatic carriers in malaria persistence and the need for targeted surveillance and control efforts. Parasites Hosts Dis. 2025;63(1):57–65.

Chacha GA, Francis F, Mandai SS, Seth MD, Madebe RA, Challe DP, et al. Prevalence and drivers of malaria infection among asymptomatic and symptomatic community members in five regions with varying transmission intensity in mainland Tanzania. Parasit Vectors. 2025;18(24):1–18. Available from: 10.1186/s13071-024-06639-1

Challe DP, Francis F, Seth MD, Tupa JB, Madebe RA, Mandara CI, et al. Prevalence and risk factors associated with infections caused by Plasmodium parasites at the micro-geographic level in three villages of Muheza district in Tanga region, north-eastern Tanzania. Malar J. 2025;24(312):1–13.

Mandai SS, Francis F, Challe DP, Seth MD, Madebe RA, Petro DA, et al. High prevalence and risk of malaria among asymptomatic individuals from villages with high prevalence of artemisinin partial resistance in Kyerwa district of Kagera region, north - western Tanzania. Malar J. 2024;23(197):1–18. Available from: 10.1186/s12936-024-05019-5

Mahama T. Bayesian Hierarchical modeling for small-area estimation of disease Burden. Int J Sci Res Arch. 2022;07(02):807–27.

Zheng W, Elliott A, Miller C, Scott M. A Bayesian INLA-SPDE Approach to Spatio-Temporal Point-Grid Fusion with Change-of-Support and Misaligned Covariates. arXiv. 2025;2511(14535v1):1–32.

TACAIDS, ZAC, NBS, OCGS, ICF. Tanzania HIV/AIDS and Malaria Indicator Survey 2011-12: Key Findings. 2013; Dar es Sal (Tanzania):1–16.

MoH, NBS, OCGS, ICF. Tanzania Demographic and Health Survey and Malaria Indicator Survey 2022 Final Report. 2023; Dodoma, Ta (Maryland, USA):1–919.

Ishengoma DS, Mmbando BP, Mandara CI, Chiduo MG, Francis F, Timiza W, et al. Trends of Plasmodium falciparum prevalence in two communities of Muheza district, North-eastern Tanzania: correlation between parasite prevalence, malaria interventions and rainfall in the context of re - emergence of malaria after two decades of progr. Malar J. 2018;17(252):1–11. Available from: 10.1186/s12936-018-2395-1

Ishengoma DS, Mmbando BP, Segeja MD, Alifrangis M, Lemnge MM, Bygbjerg IC. Declining burden of malaria over two decades in a rural community of Muheza district. Malar J. 2013;12(338):1–12. Available from: Malaria Journal

Francis F, Ishengoma DS, Mmbando BP, Rutta ASM, Malecela MN, Mayala B, et al. Deployment and use of mobile phone technology for real-time reporting of fever cases and malaria treatment failure in areas of declining malaria transmission in Muheza district, north-eastern Tanzania. Malar J. 2017;16(308):1–14.

Liheluka EA, Massawe IS, Chiduo MG, Mandara CI, Chacky F, Ndekuka L, et al. Community knowledge, attitude, practices and beliefs associated with persistence of malaria transmission in North-western and Southern regions of Tanzania. Malar J. 2023;22(304):1–16. Available from: 10.1186/s12936-023-04738-5

Ishengoma DS, Mandara CI, Bakari C, Fola AA, Madebe RA, Seth MD, et al. Evidence of artemisinin partial resistance in northwestern Tanzania: clinical and molecular markers of resistance. Lancet Infect Dis. 2024;24:1225–33. Available from: 10.1016/S1473-3099(24)00362-1

Popkin-Hall ZR, Seth MD, Madebe RA, Budodo R, Bakari C, Francis F, et al. Prevalence of non - falciparum malaria infections among asymptomatic individuals in four regions of Mainland Tanzania. Parasit Vectors. 2024;17(153):1–6. Available from: 10.1186/s13071-024-06242-4

Popkin-Hall ZR, Seth MD, Madebe RA, Budodo R, Bakari C, Francis F, et al. Malaria Species Positivity Rates Among Symptomatic Individuals Across Regions of Differing Transmission Intensities in Mainland Tanzania. J Infect Dis. 2024;229(4):959–68. Available from: 10.1093/infdis/jiad522

Bakari C, Jones S, Subramaniam G, Mandara CI, Chiduo MG, Rumisha S, et al. Community-based surveys for Plasmodium falciparum pfhrp2 and pfhrp3 gene deletions in selected regions of mainland Tanzania. Malar J. 2020;19(391):1–12. Available from: 10.1186/s12936-020-03459-3

Ishengoma DS, Francis F, Mmbando BP, Lusingu JPA, Magistrado P, Alifrangis M, et al. Accuracy of malaria rapid diagnostic tests in community studies and their impact on treatment of malaria in an area with declining malaria burden in north-eastern Tanzania. Malar J. 2011;10(176):1–13. Available from: http://www.malariajournal.com/content/10/1/176

Seth MD, Popkin-hall ZR, Madebe RA, Budodo R, Bakari C, Lyimo BM, et al. Prevalence of subpatent Plasmodium falciparum infections in regions with varying transmission intensities and implications for malaria elimination in Mainland Tanzania. Malar J. 2025;24(101):1–14.

Budodo R, Mandai SS, Bakari C, Seth MD, Francis F, Chacha GA, et al. Performance of rapid diagnostic tests, microscopy, and qPCR for detection of Plasmodium parasites among community members with or without symptoms of malaria in villages located in North - western Tanzania. Malar J. 2025;24(115):1–18. Available from: 10.1186/s12936-025-05361-2

NMCP. The United Republic of Tanzania: National Guidelines for Malaria Diagnosis, Treatment and Preventive Therapies 2020. 2020.

MoH. Standard Treatment Guidelines & National Essential Medicines List Tanzania Mainland. 2017;1–464.

Mougeni F, Lell B, Kandala NB, Chirwa T. Bayesian spatio-temporal analysis of malaria prevalence in children between 2 and 10 years of age in Gabon. Malar J. 2024;23(57):1–16. Available from: 10.1186/s12936-024-04880-8

Roques L, Allard D, Soubeyrand S. Spatial statistics and stochastic partial differential equations: a mechanistic viewpoint. arXiv. 2014;2011(05724v1):1–30.

Damien BG, Sode AI, Bocossa D, Ndille EE, Aguemon B, Corbel V, et al. Bayesian spatial modelling of malaria burden in two contrasted eco - epidemiological facies in Benin (West Africa): call for localized interventions. BMC Public Health. 2022;22(1754):1–15.

Gaedke-merzhäuser L, Krainski E. Integrated Nested Laplace Approximations for Large-Scale Spatial-Temporal Bayesian Modeling. arXiv. 2023;2303(15254v1):1–22.

Bivand RS, Virgilio GR, Rue H. Spatial Data Analysis with R - INLA with Some Extensions. J Stat Softw. 2015;63(20):1–31.

Du H, Keller B, Alacam E, Enders C. Comparing DIC and WAIC for multilevel models with missing data. Behav Res Methods. 2024;56:2731–50.

Zong L, Ngarukiyimana JP, Yang Y, Yim SHL, Zhou Y, Wang M, et al. Malaria transmission risk is projected to increase in the highlands of Western and Northern Rwanda. Common Earth Environ. 2024;5(559):1–9. Available from: 10.1038/s43247-024-01717-9

Mayilsamy M, Parthasarathy R, Veeramanoharan R, Rajaiah P. Impact of climatic factors on the occurrence of malaria in hyper, high, moderate and low endemic States in India from 1995 to 2023. Malar J. 2025;24(113):1–11. Available from: 10.1186/s12936-025-05326-5

Nyasa RB, Awatboh F, Kwenti TE, Titanji VPK, Ayamba NLM. The effect of climatic factors on the number of malaria cases in an inland and a coastal setting from 2011 to 2017 in the equatorial rain forest of Cameroon. BMC Infect Dis. 2022;22(461):1–12. Available from: 10.1186/s12879-022-07445-9

Kahamba NF, Okumu FO, Jumanne M, Kifungo K, Odero JO, Baldini F, et al. Geospatial modelling of dry season habitats of the malaria vector, Anopheles funestus, in south-eastern Tanzania. Parasit Vectors. 2024;17(38):1–14. Available from: 10.1186/s13071-024-06119-6

Dzokoto MK, Mensah BT, Osei-owusu W, Adam I, Kwashie SDY. The influence of enhanced vegetation index, proximity to national borders, proximity to protected areas, and proximity to water on malaria case prevalence in Sub-Saharan Africa, 2000-2020. Int J Community Med Public Heal. 2023;10(10):3506–16.

Dida GO, Anyona DN, Abuom PO, Akoko D, Adoka SO, Matano A said, et al. Spatial distribution and habitat characterization of mosquito species during the dry season along the Mara River and its tributaries, in Kenya and Tanzania. Infect Dis Poverty. 2018;7(2):1–16.

Otambo WO, Onyango PO, Wang C, Olumeh J, Ondeto BM, Lee MC, et al. Influence of landscape heterogeneity on entomological and parasitological indices of malaria in Kisumu, Western Kenya. Parasit Vectors. 2022;15(340):1–13. Available from: 10.1186/s13071-022-05447-9

Palaniyandi M. The Environmental Risk Factors Significant to Anopheles Species Vector Mosquito Profusion, P. falciparum, P. vivax Parasite Development, and Malaria Transmission, Using Remote Sensing and GIS: Review Article. Indian J Public Heal Res. 2021;12(4):162–71.

Gbaguidi GJ, Topanou N, Filho WL, Begedou K, Ketoh GK. Environmental and socio-economic determinants of malaria transmission in West Africa: a systematic review. One Heal Outlook. 2025;7(47):1–16.

Iddrisu A Karim, Otoo D, Hinneh G, Kanyiri YD, Samuel KY, Kubio C, et al. Identifying Malaria Hotspot Regions in Ghana Using Bayesian Spatial and Spatiotemporal Models. Infect Dis Immun. 2024;4(2):69–78.

Monteiro GM, Aïkpon RY, Dandonougbo C, Sedda L, Djogbenou LS. Identification of malaria hotspots in southwestern Benin through spatial joint modelling of malaria incidence and vector abundance. VeriXiv. 2025;2(273):1–20.

Clarotto L, Allard D, Romary T, Desassis N. The SPDE approach for spatio-temporal datasets with advection and diffusion. HAL open Sci. 2024;03762403v2:1–35.

Stresman G, Bousema T, Cook J. Malaria Hotspots: Is There Epidemiological Evidence for Fine-Scale Spatial Targeting of Interventions? Trends Parasitol. 2019;35(10):822–34. Available from: 10.1016/j.pt.2019.07.013

Bonza Z, Katapa R, Msengwa A. Bayesian predictive modelling to ascertain factors affecting cattle milk production in Tanzania: Evidence from the national panel surveys 2012 – 2021. Vet Anim Sci. 2025;27(100404):1–7. Available from: 10.1016/j.vas.2024.100404

Alahmadi H, Moraga P. Bayesian modelling for the integration of spatially misaligned health and environmental data. Stoch Environ Res Risk Assess. 2025;39:1485–99.

Anderson MJ, Walsh DCI, Sweatman WL, Punnett AJ. linear models of species’ responses to environmental and spatial gradients. Ecol Lett. 2022;25(2739):2739–52.

Msugupakulya BJ, Mhumbira NS, Mziray DT, Kilalangongono M, Jumanne M, Ngowo HS, et al. Field surveys in rural Tanzania reveal key opportunities for targeted larval source management and species sanitation to control malaria in areas dominated by Anopheles funestus. Malar J. 2024;23(344):1–19. Available from: 10.1186/s12936-024-05172-x.

Oyibo W, Ntadom G, Uhomoibhi P, Oresanya O, Ogbulafor N, Ajumobi O, et al. Geographical and temporal variation in reduction of malaria infection among children under 5 years of age throughout Nigeria. BMJ Glob Heal. 2021;6(e004250):1–10.

Mwangungulu SP, Dorothea D, Ngereja ZR, Kaindoa EW. Geospatial-based model for malaria risk prediction in Kilombero valley, South-eastern Tanzania. PLoS One. 2023;18(10):1–23. Available from: 10.1371/journal.pone.0293201

Wanjala CL, Kweka EJ. Impact of Highland Topography Changes on Exposure to Malaria Vectors and Immunity in Western Kenya. Front Public Heal. 2016;4(227):1–11.

Borkovska O, Pollard D, Hamainza B, Kooma E, Renn S, Schmidt J, et al. Developing High-Resolution Population and Settlement Data for Impactful Malaria Interventions in Zambia. J Environ Public Health. 2022;2022(2941013):1–8.

Nkya TE. Insecticide Resistance Management in Malaria Vectors in Tanzania Mainland (2015 – 2024): A Scoping Review. Trop Med $ Int Heal. 2025;0:1–14.

Kelly GC, Seng CM, Donald W, Taleo G, Nausien J, Iata H, et al. A spatial decision support system for guiding focal indoor residual spraying interventions in a malaria elimination zone. Geospat Health. 2011;6(1):21–31.

